# Standardized incidence ratio of the COVID-19 pandemic: a case study in a Midwestern state

**DOI:** 10.1101/2021.09.28.21263671

**Authors:** Emma Spors, Semhar Michael

**Affiliations:** Mathematics and Statistics, South Dakota State University, 905 Campanile Ave, Brookings, SD, USA

**Keywords:** COVID-19, Standardized Incidence Ratio, Lasso regression, Generalized Linear Models, age-adjustment

## Abstract

**Motivation:** The Coronavirus disease 2019 (COVID-19) has made a dramatic impact around the world, with some communities facing harsher outcomes than others. We sought to understand how counties in the state of South Dakota (SD) fared compared to expected based on a reference population and what factors contributed to negative outcomes from the pandemic in SD.

**Methods:** The Standardized Incidence Ratios (SIR) of all counties, using age-adjusted and crude adjusted hospitalization and death rates were computed using the SD age-adjusted rate as a reference population. In addition, a penalized generalized linear regression model was used to identify factors that are associated with COVID-19 hospitalization and death rates. This model was then used to compute a new SIR after controlling for other socio-demographic and -economic factors.

**Results:** We identified counties that had more or less severe outcomes than what would be expected based on the rate of SD after age adjustment. Additionally, race, education, and testing rate were some of the significant factors associated with the outcome. The SIR values after controlling for these additional factors showed change in magnitude from the range of 4 times more severe to 1.5 times more severe outcome than what is expected. Interestingly the lower end of this interval did not have a major change.

**Conclusion:** The age adjusted SIR model used in this study allowed for the identification of counties with more or less severe than what is expected based on the state rate. These counties tended to be those with high nonwhite percentage, which mostly included counties with American Indian reservations. Although several predictors are associated with hospitalization and deaths, the penalized model confirmed what is already reported in literature that race and education level have a very high association with the outcome variables. As can be expected the further adjusted SIR mostly changed in those counties with higher than expected outcomes. We believe that these results may provide useful information to improve the implementation of mitigation strategies to curb the damage of this or future pandemics by providing a way for data-driven resource allocation.

## 1. Background and motivation

COVID-19 is the respiratory disease caused by “severe acute respiratory syndrome coronavirus 2” (SARS-CoV-2), a novel coronavirus that appeared in 2019. Officially declared a global pandemic on March 11, 2020 by the World Health Organization, COVID-19 has made a drastic and lasting impact on the world [1].This study sought to understand how the counties fared relative to each other and what socio-economic and -demographic factors contributed to COVID-19 hospitalizations and deaths in SD’s counties. While COVID-19 infections may be mild, some cases cause severe illness which includes hospitalization, intensive care, the need for artificial ventilation, and possibly death. Persons with certain conditions are more likely to become severely affected by COVID-19 than others. Most notably are older adults, as 80 percent of COVID-19 deaths occur in people over the age of 65 [2]. In addition, those with long-standing systemic health and social inequalities, such as many racial and ethnic minority groups and people with disabilities are at an increased risk [11]. Specific medical conditions that are also associated with higher risk include, but are not limited to (in alphabetical order): cancer, chronic kidney disease, chronic lung diseases, neurological conditions, diabetes, heart conditions, immunocompromised state, liver disease, over weight or obesity, pregnancy, and sickle cell disease [11]. In addition, life style factors, such as smoking and substance abuse, also may place people at increased risk [11]. The COVID-19 pandemic has led to a huge effort in scientific research with a variety of different approaches and methodology. Many researchers have used bioinformatics to understand the disease from a a clinical perspective, including sequencing the genome of the virus and documenting the physiological response within individuals responding the infection *citations*. Additional work has gone into rigorously studying individual risk factors associated with severe infection as well as using statistical forecasting models to predict the magnitude of future cases, hospitalizations, and deaths. A subset of available literature has examined some county-level socio-demographic and -economic factors as predictors of the pandemic. One study used penalized regression analysis to identify those factors that were associated with increased COVID-19 cumulative case rates in the state of Georgia [21]. Richmond et. all considered a different pool of socioeconomic and demographic that what was considered for this study, and found that ??

There were several outcomes that could be measured from the COVID-19 pandemic. In this paper COVID-19 hospitalizations and deaths at the county level were considered as response variables. The standardized incidence ratios before age adjustment, after age adjustment, and after adjusting for other socio-demographic and -economic factors and their corresponding confidence bands are computed. In addition, several social, economic, and demographic variables, that have been reported in the literature as risk factors for infectious disease transmission, were also considered at the county level. We considered penalized generalized linear models to identify factors that are associated to disease spread [15].

The paper is organized as follows: In Section 2 we provide the data sources and statistical methods used for data analysis. In Section 3 the results of the analysis are presented. Section 4 discusses our main findings and the corresponding literature. Concluding remarks are given in Section 5.

## 2. Methods

### 2.1. Data sources

The South Dakota (SD) COVID-19 data were received directly from the SD Department of Health (SDDOH) [23]. This study specifically examined reported COVID-19 cases, hospitalizations, and deaths at a county level. Based on definitions from the SDDOH, cases included persons who met the national surveillance definition for COVID-19. A person who had a positive PCR test for SARS-CoV-2 was a confirmed case and a person with a positive antigen test was a probable case. A person was listed as hospitalized if, at any point during the infection, they were hospitalized under transmission-based precautions. Deaths reflected the number of people who died and COVID-19 was listed as the cause of death, or significant contributor to death as determined by the judgement of the healthcare provider or coroners completing the death certificate [3].

The dataset obtained from SDDOH included information on individual cases of COVID-19 in SD. Details encompassed the date the positive test was reported to the SDDOH, a positive PCR test indicator (yes/no), positive antigen test indicator (yes/no), recovery date, current county of residence, age category, hospitalization or death indicators (yes/no). If necessary, date of hospitalization and discharge and date of death were included [23]. The age category was represented by numbers one through nine, which corresponded to ten-year age groupings including 0 to 9, 10 to 19, 20 to 29, 30 to 39, 40 to 49, 50 to 59, 60 to 69, 70 to 79 years of age, and 80 years of age and older. An additional dataset containing the daily count of PCR and antigen COVID-19 tests per county was provided[24]. The testing data were not broken down by age group.

We selected the following socioeconomic and demographic factors at the county level for analysis: nonwhite percentage, educational attainment (percentage of population with a bachelor’s degree or higher), percentage of population with total annual income at or below a specified poverty threshold (determined by household size and composition) as designated by the U.S. Census Bureau, median income, unemployment percentage, uninsured percentage, cardiovascular disease hospitalization rate per 1,000 population, diabetes percentage (Type I & II), obesity percentage defined by BMI, physical inactivity percentage, and rate of providers,. The source and description of each factor are described in detail in the appendix (See Section 6).

The initial dataset had several incongruities noted for hospitalization dates. For example, there were dates of hospitalization that were before the pandemic officially reached SD. Data cleaning was performed by changing the listed date to a more appropriate date based on available information from the specific case. For example, the date of hospitalization listed 01/20 was changed to 01/21 based on the date a positive test result was reported to SDDOH.

### 2.2. Data Preparation and Adjustments

After data cleaning, the data for cases, hospitalization, and death were transformed from information on an individual level to counts by county of residence per age group (1 through 9), by day (from Mar 10, 2020 through May 1, 2021). One of the common practices in accounting for population is to calculate crude rates (*i.e*. the counts (of hospitalization or death) per county will be divided by the population of the county and multiplied by a number such as 100,000 to find he crude rate for a county per 100,000). In this work, one of the adjustments done to the data is to compute the crude death and hospitalization rates per 100,000 by county. In addition, it is well known that severe illness from COVID-19, in the time frame this report covers (before the delta variant), disproportionately impacts older individuals. The age distribution for SD and two counties is shown in Figure 1. For example, In college towns such as Brookings, the age distribution (based on the 2019 population estimates [27]) is different than SD age distribution and many of the other counties in SD (see Figure 1). The Brookings County has a younger population than both Brown County and SD overall. Thus, crude rates of hospitalization or deaths per 100,000 may be less in Brookings County compared to counties with an older population simply due to age differences. To account for this difference, the age-adjusted rate per 100,000 is standardized to the values across counties with respect to their age distribution. The daily age-adjusted rate per 100, 000, *R*, was calculated as

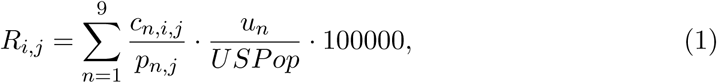

where *i* increments daily for the dates from 10-March-2020 to 5-May-2021, *j* = 1, …, 66 is a county in SD, *n* = 1, …, 9 represents a specific age group, *c*_*n,i,j*_ represents the count of a particular outcome (hospitalization or death) for age group *n* on day *i* in county *j, p*_*n,j*_ is the population size for the *n*th age group in the *j*th county, *USPOP* represents the United States (US) population based on the 2019 Census estimates and *u*_*n*_ is the US population in the age group *n*. The daily age-adjusted rates per 100, 000 were calculated for hospitalizations and deaths. Since the testing data was not broken down by age group, we calculated the daily crude testing rate per 100,000 as 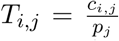 · 100000, where *c*_*i,j*_ is the count of tests reported on *i*th day from *j*th county and *p*_*j*_ is the population of the *j*th county.

**Figure 1.:**
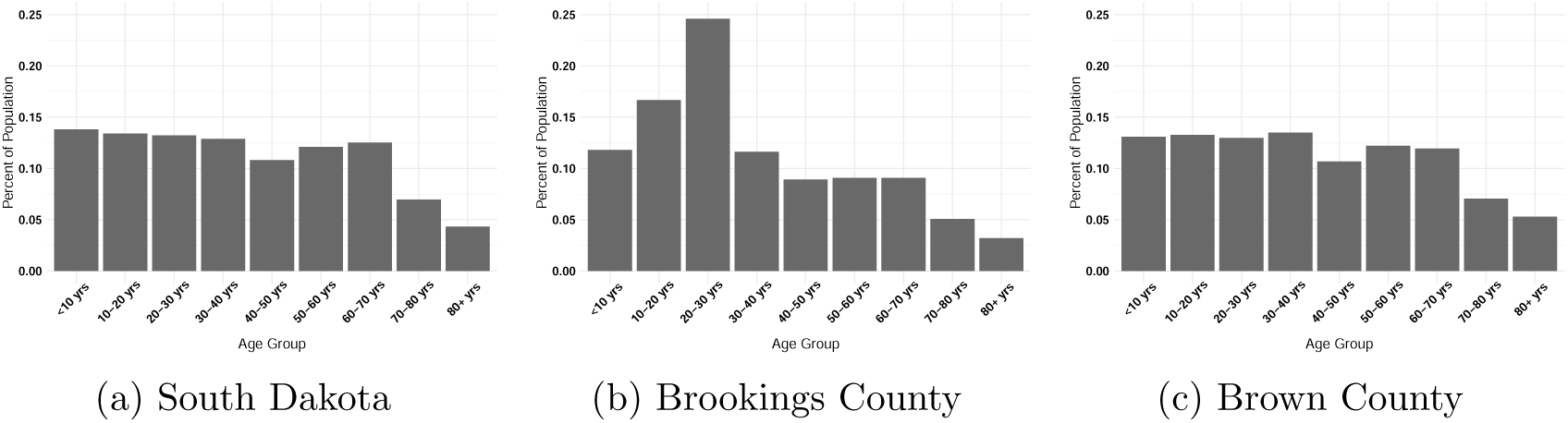
Age distribution for South Dakota, Brookings County, and Brown County.

For SIR values and linear models, cross-sectional data are needed. Therefore, cumulative counts at a specific date were considered. Similar to daily counts, the cumulative crude and age-adjusted rate per 100,000 was also calculated for deaths and hospitalizations. For age adjusted cumulative, the following steps were followed. After finding the cumulative hospitalizations and deaths for each county up to May 1, 2021, the cumulative age adjusted rate was calculated with 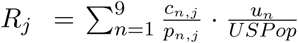 · 100000, where *c*_*n,j*_, *p*_*n,j*_, and *u*_*n*_ are defined similarly as in Equation 1. Again, the testing was calculated by cumulative tests per 100,000 with 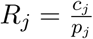 · 100000.

### 2.3. Penalized Generalized Linear Model

To understand how socioeconomic factors impacted COVID-19 hospitalizations and deaths, we utilized generalized linear models (GLM) with Poisson and Gaussian link functions. In general, given the response variable *Y*_1_, …, *Y*_*n*_ and explanatory variables ***X***_**1**_, …, ***X***_***n***_, assume *μ*_*i*_ = *E*(*Y*_*i*_), the GLM has the following structure, *g*(*μ*_*i*_) = ***X***_***i***_*β*, and *g* is called the link function. In GLM we assume that *Y*_*i*_ follows an exponential family distribution such as the Gaussian, Binomial, Poisson, etc [31]. The *β*_0_, *β*_1_, *β*_2_, …, *β*_*p*_ are estimated using the maximum likelihood estimation approach. For this method, the coefficients that maximize the likelihood function are obtained. In our data, however, since there were a large number of potential predictors and several of them exhibited co-linearity (see Figure 5), we chose to use the penalized GLM for feature selection. Specifically, the Lasso GLM incorporates 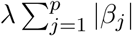 as a penalty in the likelihood function[14, 31]. Lasso regression will force some of the coefficient estimates to be exactly zero as *λ* increases, effectively dropping the variables from the model. In addition to identifying significant factors and their relationship to the response, the results of this model yields the expected value of the response after controlling for the factors considered in the model.

### 2.4. Standardized Incidence Ratio

A Standardized incidence ratio (SIR) was used to compare how counties fared in terms of cumulative hospitalizations and deaths. This was done by comparing the observed value in a county to an expected value and was calculated with 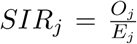, where *O*_*j*_ was the observed value from the *j*th county and *E*_*j*_ was the expected value for the *j*th county. The confidence intervals for the *SIR* indices were calculated with 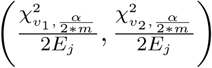, where *v*_1_ = 2*O*_*j*_, *v*_2_ = 2(*O*_*j*_ + 1) and 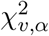 was from the Chi-Squared distribution with the critical value at *α* = 0.05 and *v* degrees of freedom [19]. To adjust for multiple comparisons, we used a Bonferroni correction [4, 9] and divided the critical value by 2 · *m*, where *m* = 66 was the number of comparisons to be made between counties.

An SIR value was calculated for the adjusted cumulative hospitalizations and deaths for all counties in SD. To compute the observed and expected adjusted cumulative rates, we considered three rates. These were the crude cumulative rate, the age adjusted cumulative rate, and expected cumulative rate obtained from the penalized GLM model after controlling for other significant factors. Therefore, the first indicates SIR using crude rates, then SIR’s after adjusting for age distribution of the counties, and lastly SIR’s with age adjustment and controlling for other factors. Note that in all three cases values greater than one indicate that that county was experiencing a higher rate of that measure. Conversely, if the value was lower than one, then that county was experiencing a lower rate of that measure relative to what was expected. If the confidence interval did not include 1 this indicates that the county was significantly different than what was expected. For ease of presentation and interpretation of the results we present *SIR* − 1 values. Hence, the values below zero indicate that the observed values are less than the expected and counties that had more than expected will be above zero.The confidence intervals will be shifted in a similar manner.

All data preparation and statistical analysis were performed using R and RStudio [20, 22]. The package *tidyverse* was used for general data manipulation and all graphs were created with the *ggplot2* package [29, 30]. Lasso regression was completed with the R package *glmnet* [14].

## 3. Results

### 3.1. Exploratory data analysis

To better understand the data, several techniques were used to complete exploratory data analysis (EDA). All our analysis in the EDA section will use the age-adjusted rates per 100,000 for cases, hospitalization, and deaths and crude rates for testing data. We first explored the trends in the COVID-19 data. The daily age-adjusted cases of the counties in SD are shown in Figure 2. In this plot we can see the major outbreak (peak) at around Nov-Dec 2020. We also note the initial surge in Minnehaha County, where an outbreak at a pork processing plant (Smithfield Foods Incorporation) occurred[26]. The two highest daily rates corresponded to the mass testing at the facility in … County []. The map of South Dakota with age-adjusted cumulative hospitalizations and deaths are shown in Figure 3. As expected, we see a strong association between hospitalization and death rate among the counties. Also note that the west-central part of the state has higher rates of hospitalizations and deaths. Figure 4 shows the cumulative age-adjusted hospitalizations and deaths SD counties. For hospitalization, there seems to be two groups with light gray and darker gray shades in the plot. For deaths, we can see three groups of counties with those separated at the top with like grey shade, those in the middle with darker grey, and another group with close to zero deaths at the bottom with light grey shades.

**Figure 2.:**
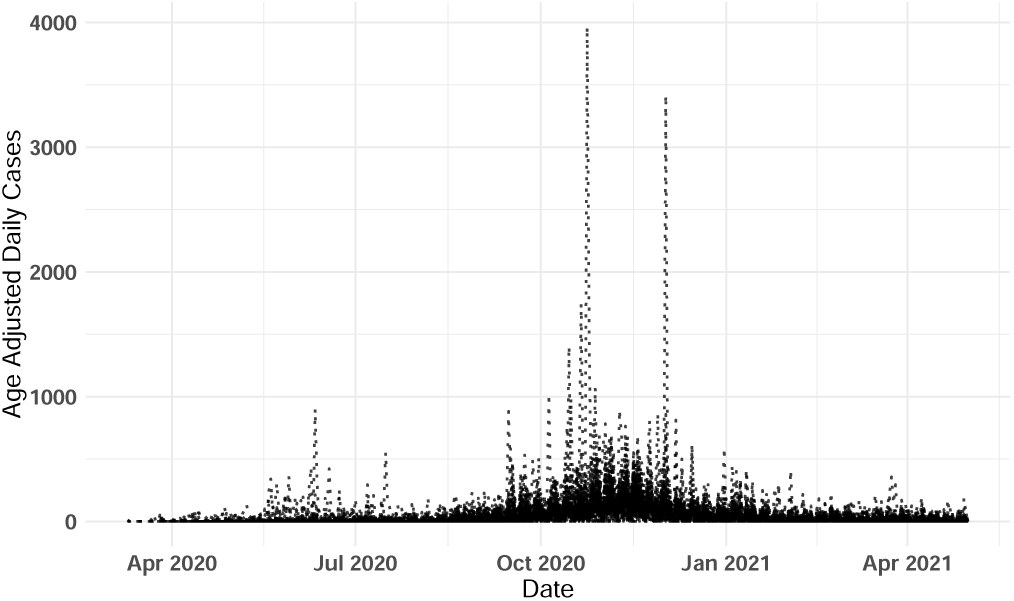
Daily age-adjusted cases per 100,000 for the top 10 counties.

**Figure 3.:**
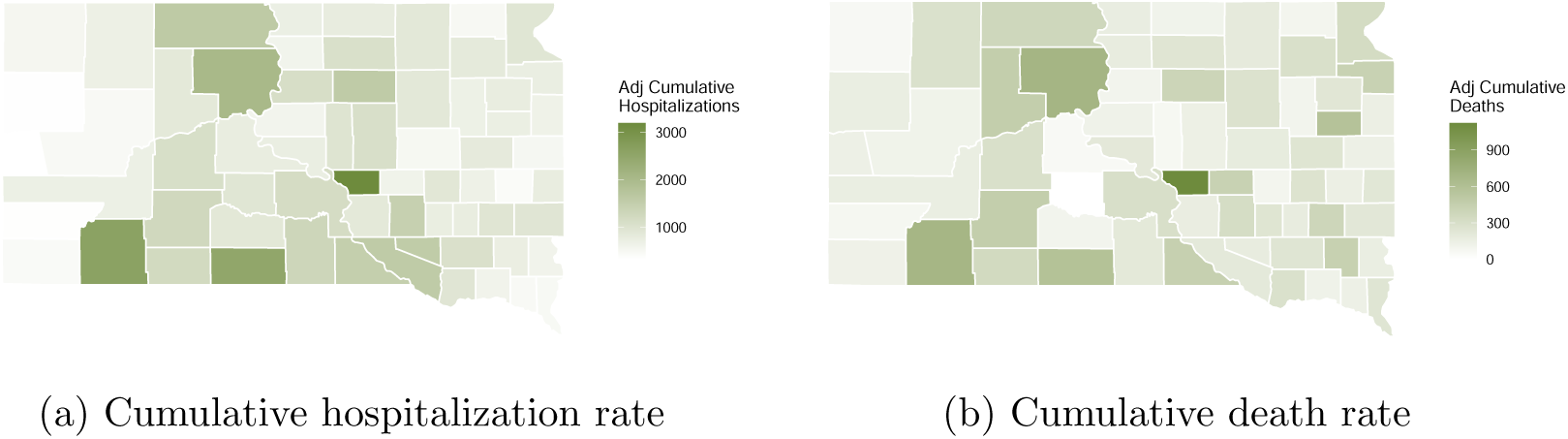
Age-adjusted rates as of May 1st, 2021 in South Dakota.

**Figure 4.:**
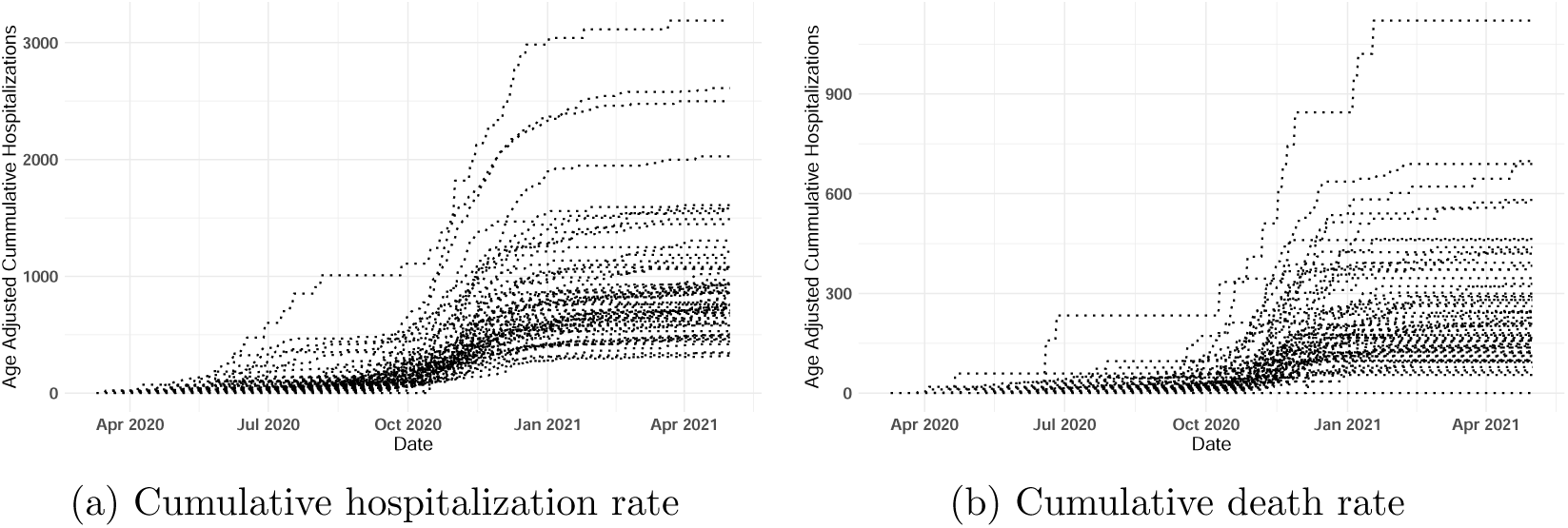
Trend of cumulative age-adjusted rates per 100,000 in South Dakota.

The numerical summaries of the demographic and socioeconomic explanatory variables are given in Table 1.A more detailed graphical summary of the socioeconomic explanatory variables are given in Appendix 6. Figure 5 shows the correlation between the explanatory variables. It can be observed that most of the variables have high correlation, explaining the need for feature selection methods. For example, as expected, median income is highly correlated with education (0.6) and inversely correlated with poverty and uninsured rates (−0.7 and −0.6, respectively).

**Table 1.:** Numerical summary of the demographic and socioeconomic factors considered in this study.

|  | Minimum | Q1 | Median | Q3 | Maximum |
| --- | --- | --- | --- | --- | --- |
| Non white | 2.13 | 3.99 | 7.24 | 16.23 | 94.65 |
| Bachelors degree and above | 8.50 | 18.73 | 22.00 | 26.23 | 49.90 |
| Poverty | 4.00 | 8.53 | 11.15 | 14.7 | 55.50 |
| Median income | 24331 | 49811 | 55980 | 54279 | 82473 |
| Unemployment | 2.30 | 2.80 | 3.00 | 3.90 | 8.90 |
| Uninsured | 6.40 | 10.43 | 11.65 | 15.50 | 21.90 |
| CVD hospitalizations | 22.10 | 36.35 | 42.95 | 49.48 | 86.70 |
| Obesity | 22.20 | 29.45 | 33.10 | 39.05 | 52.00 |
| Physical inactivity | 15.80 | 20.05 | 21.75 | 25.38 | 37.10 |
| Number of providers | 0.00 | 21.81 | 61.27 | 125.51 | 671.60 |

**Table 2.:** Linear regression models for cumulative age adjusted, case, hospitalizations, and death rate with most socioeconomic factors.

| | Coefficient | $t$ Value | $P(> \vert t\vert )$ | Coefficient | $t$ Value | $P(> \vert t\vert )$ |
| --- | --- | --- | --- | --- | --- | --- |
| Intercept | 887.90 | 1.72 | 0.089 | 114.50 | 0.57 | 0.57 |
| Nonwhite percentage | 12.38 | 3.67 | 0.00056 | 2.25 | 1.71 | 0.093 |
| Educational Attainment | -17.70 | -2.021 | 0.048 | -6.17 | -1.87 | 0.066 |
| CVD Hospitalizations | 2.98 | 0.56 | 0.58 | 0.87 | 0.42 | 0.68 |
| Diabetes (Type I & II) | 7.77 | 0.47 | 0.64 | 10.46 | 1.62 | 0.11 |
| Obesity Percentage | -6.95 | 0.90 | -0.51 | 3.45 | 0.91 | 0.37 |
| Physical Inactivity | -2.57 | -0.71 | 0.48 | -4.19 | -0.81 | 0.42 |
| Number of providers | -0.005 | -0.013 | 0.99 | 0.10 | 0.64 | 0.53 |
| Cumulative Tests | 0.0023 | 2.42 | 0.019 | 0.00052 | 1.402 | 0.17 |

**Table 3.:** Penalized regression models for cumulative age adjusted case, hospitalizations, and death rates selected demographic and socioeconomic factors.

| | Coefficient | $t$ Value | $P(> \vert t\vert )$ | Coefficient | $t$ Value | $P(> \vert t\vert )$ |
| --- | --- | --- | --- | --- | --- | --- |
| Intercept | 1160.53 | 5.73 | $2.99 \cdot 10^{-07}$ | 167.08 | 7.49 | $2.55 \cdot 10^{-10}$ |
| Nonwhite percentage | 12.49 | 5.91 | $1.49 \cdot 10^{-07}$ | 4.69 | 6.51 | $1.36 \cdot 10^{-08}$ |
| Educational Attainment | -19.37 | 7.49 | 0.012 | - | - | - |

**Figure 5.:**
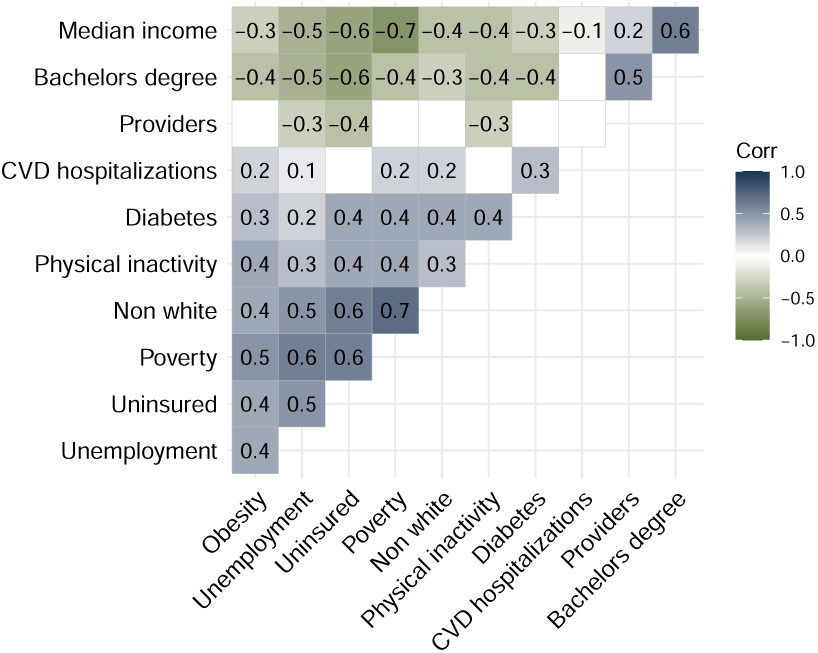
Correlation plot for some of the socioeconomic factors.

### 3.2. SIR and Penalized GLM

#### Results

A GLM model with a set of variables without some of the highly correlated variables and a penalized Poisson regression with lasso penalty were fitted for age-adjusted cumulative hospitalization and death rates on May 1, 2021. The lambda for the Lasso penalty was chosen based on cross-validation method. For cumulative hospitalizations, the lasso model selected the nonwhite percentage and educational attainment as important variables. After creating the linear model, there was a positive association between the cumulative rate of hospitalizations and the nonwhite percentage. Conversely, there was a negative association between the cumulative rate of hospitalization and educational attainment. That is, a county with a higher number of college-educated residents saw fewer hospitalizations. For cumulative deaths, lasso regression selected nonwhite percentage. The linear model revealed that there was a positive association between cumulative deaths and the nonwhite percentage.

After fitting the penalized GLM models to the data we computed the expected cumulative hospitalization and death rate for each county. This then was used to compute new predicted SIR values after controlling for race and educational attainment in case of hospitalization and race in case of deaths. The results of both SIR’s with just age adjustment and with both age and other factor adjustment are presented in Figures 6 and 7. For age-adjusted rates of hospitalization, from the top ten counties in SD, Lawrence, Clay, Beadle, and Brookings had the lowest significant *SIR* values with Lawrence County experiencing about 60 percent fewer hospitalizations and Brookings experiencing about 36 percent fewer hospitalizations than expected. Minnehaha had 17 percent more and Brown had 10 percent more cases than expected. For deaths, from the top ten counties in SD, Yankton, Brookings, Clay, and Lawrence had the lowest *SIR* values with Yankton experiencing about 43 percent fewer deaths and the others about 35 percent fewer deaths. We note that similar counties tend to have more than expected on severe outcomes.

**Figure 6.:**
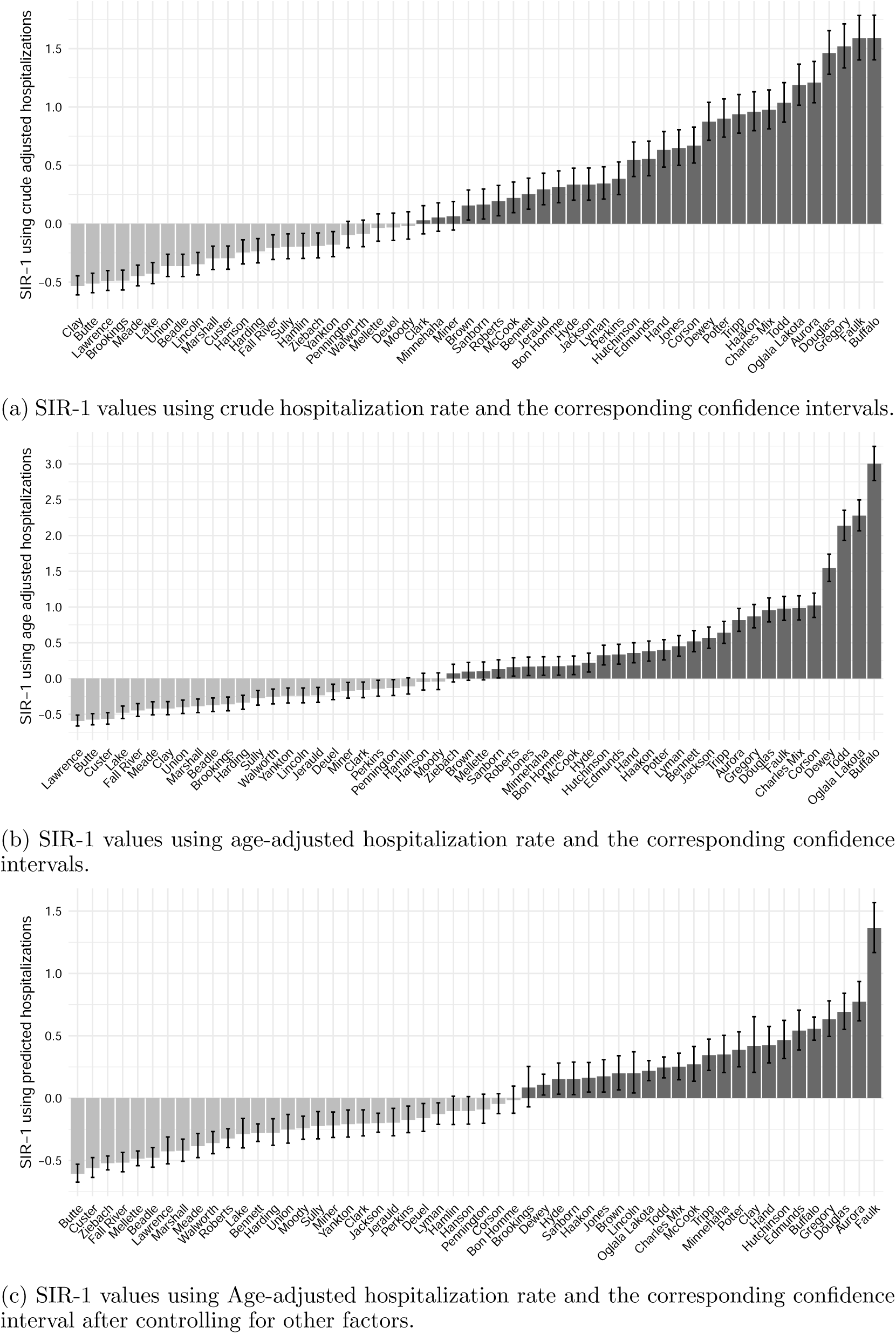
SIR-1 values for cumulative hospitalization rates as of May 1st, 2021 for counties in South Dakota with at least one significant SIR-1 value.

**Figure 7.:**
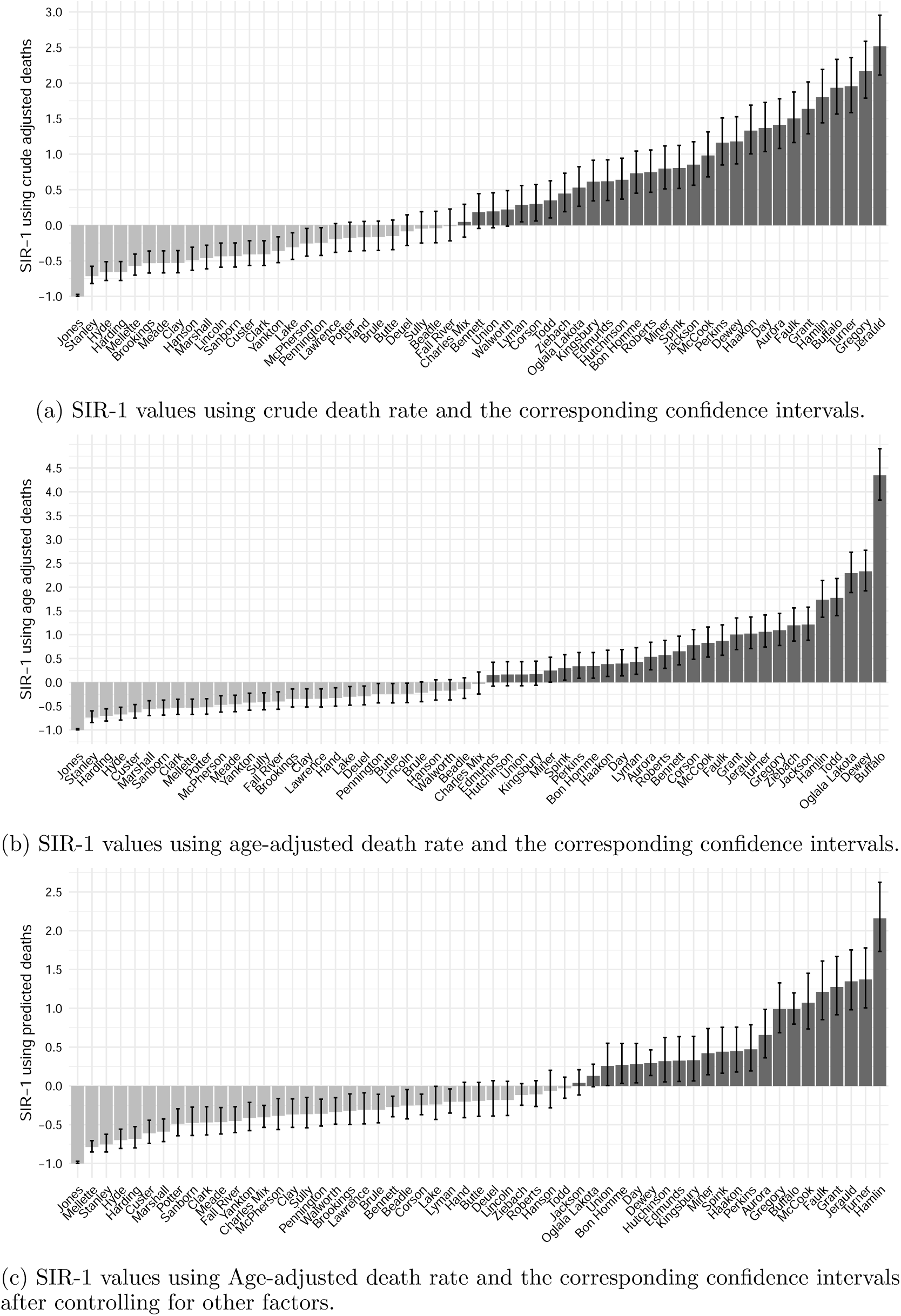
SIR-1 values for cumulative death rates as of May 1st, 2021 for counties in South Dakota with at least one significant SIR-1 value.

After controlling for other factors using the GLM model, the range of SIR values for counties with significant SIR’s shifted from a maximum of around 3 to 1.25 for hospitalization rate and from around 4.5 to 2.5 in the case of death rate. Especially, counties with a large non-white population have moved towards the middle of the SIR graph or have become non-significant and have been dropped.

## 4. Discussion

The SIR model shows how different parts of SD responded to the pandemic. by demonstrating the spatial patterns and identifies the counties that had more share in the rate of severe outcomes and those that had less. We noted that in the top 11 counties, most counties had lower values than expected when compared to South Dakota as a baseline. Consistently, Lawrence, Brookings, and Clay had significantly lower rates of severe outcomes than expected. In contrast, Minnehaha and Davison experienced significantly higher rates of severe outcomes than expected. Looking at specific states at the worse end of the map, Dewey, Todd, Oglala Lakota, and Buffalo had twice to four times more hospitalizations and deaths that what might be expected. We note that these counties are where some of the American Indian reservations are located in SD.

Throughout all the models, the nonwhite percentage was repeatedly selected as positively associated with hospitalizations and deaths. Based on the available literature, this is not surprising. Historical treatment and continuing racial differences have contributed to large disparities between minority and white populations in the United States. In South Dakota, American Indian and Alaska Native (AI/AN) is the largest minority at 9 percent followed by two or more Races (2.5 percent) and Black or African American (2.3 percent) [27]. According to the CDC, the ratio of AI/AN to white non-Hispanic persons was 3.4 for hospitalizations and 2.0 for deaths. For Black or African Americans, it was 2.8 for hospitalization and 2.0 for deaths. That is, for all measures, both AI/AN and Black persons experienced higher rates that their white counterparts. In addition, AI/AN had the highest ratios among all minorities for hospitalizations and deaths [12]. One study that confirmed the disproportionate burden of cases on AI/AN and cited reliance on shared transportation and household size may have been a few of several factors that increased community transmission and which may have incurred higher rates of hospitalizations and deaths [16]. In addition, Indian Health Services reports that AI/AN have consistently experienced a decreased health status compared to other Americans [18]. AI/AN disproportionately suffer from high blood pressure, chronic liver disease, obesity, diabetes, heart disease, chronic lower respiratory disease among others [18]. These all contribute to an increased likelihood of hospitalization or death from a COVID-19 infection.

In the cumulative hospitalizations linear model, in addition, educational attainment was negatively associated with hospitalizations. Counties with more adults who had achieved a bachelor degree or higher had fewer COVID-19 hospitalizations. Current literature proposes two explanations. Low educational attainment (such as a high school degree or less) has been associated with adverse health effects such as coronary artery disease[32]. Scientists are now understanding the overall relationship between education and health, finding that more schooling is linked to better health and longer life [32]. Adults with more years of education tend to have access to higher paying, more stable jobs where they can accumulate wealth to invest in their health. In addition, individuals with fewer years of education tend to have more chronic health conditions, smoke, have a less healthy diet, and lack adequate exercise [32]. All of these factors can culminate in someone who is more likely to require hospitalization due to COVID-19 based on known risk factors [11]. In addition, research has shown that the pandemic has disproportionately impacted those with lower educational levels [13].

Some limitations to this study comes from the data itself. Many of the socioeconomic data sets were based on self-reported information which may confer bias to those numbers. In addition, South Dakota has a rural state with many counties being sparsely populated as the median county population was 5,430. This lead to some disproportionate rates. Take, for example, the cumulative age-adjusted death rates in Jones and Buffalo Counties. Jones had a population near 900 and reported no deaths as of May 1st. Buffalo had a population near 1,962 and reported 13 deaths. Jones County had the lowest age-adjusted death rate while Buffalo County had the highest death rate. Due to their small size, for some counties, a small difference in numbers can lead to a big change in the adjusted rates. In addition, for the SIR models, the data was used twice.

## 5. Conclusion

This study used SIR and GLM models to compare age-adjusted COVID-19 hospitalization and death rates among SD counties before and after adjusting for demographic and socioeconomic factors. To account for age difference and population in the counties age-adjusted rates were used.

For further research we suggest looking into COVID-19 positivity rates and hospitalization rates to determine if certain counties were under testing their population. This would help identify if there were likely undocumented cases in the state. In addition, time series models could be developed and used to predict future levels of the pandemic. In addition, a more flexible model that accounts for the heterogeneity in the counties of the state can be considered. Other mobility or social media related data can be incorporated to capture more variability in the response.

## Data Availability

The South Dakota Department of Health provided the data per the request of a researcher.

## Acknowledgement

We acknowledge the support of the South Dakota Department of Health in providing the COVID-19 data.This work is partially supported by the South Dakota State University’s Presidential Research Project (PREP-21, SDSU). Additional expertise was provided by Bonny Specker, PhD, Professor Emerita from South Dakota State University. The contents are solely the responsibility of the authors. No copyrighted figures, surveys, instruments, or tools were used in this study.

## 6. Appendix

### 6.1. Sources of data

1. *The US Census* provided demographic information on population estimates broken down by race and age groups in 2019. Due to South Dakota’s predominately white population (with the largest minority being Native American or Alaska Native at 9 percent), the percentage of nonwhite individuals overall was calculated instead of being broken down by racial groups. The age group demographics were necessary to compute age adjusted rates of COVID-19 cases, hospitalizations, and deaths [27].
2. *The American Community Survey* via the United States Census Bureau provided information on education attainment percentages, poverty percentages, and median income per county in SD in 2019 [5–7]. The American Community Survey is an ongoing survey that gathers information from the United States public for government and public use. For this study, education attainment was measured as the percentage of a county’s residents, aged 25 and older, who had completed a bachelor’s degree, master’s degree, doctoral degree, or professional degree [6]. The poverty statistic measured the percentage of county residents for whom poverty status was determined. The poverty threshold is adjusted based on family size and composition. It is not adjust geographically, but the thresholds are updated for inflation using the Consumer Price Index. If a family’s combined pretax income does not meet the threshold, then the whole family is considered in poverty [7]. Median income represented the median income of households in the last 12 month to 2019 inflation-adjusted dollars [5].
3. The unemployment percentage per county in 2019 for South Dakota was taken from the *Local Area Unemployment Statistics* and the South Dakota Department of Labor [25].
4. *The Small Area Health Insurance Estimates (SAHIE)* program provided estimates of health insurance coverage for residents in SD counties under the age of 65 for 2019 [8]. For an individual to be insured, they must have been currently covered by insurance from a current or former employer or union, had insurance purchased directly from an insurance company, had Medicare, been on any kind of government-assistance plan for those with low-income or disability, military health care, VA health care, and/or Indian Health Services (IHS). However, those only covered with IHS were considered uninsured because IHS was not comprehensive coverage [8]. This study specifically looked at the percentage of uninsured individuals living in each county.
5. The rate of cardiovascular hospitalizations was from the *Centers for Disease Control and Prevention’*s Division for Heart Disease and Stroke Prevention. These values represented the age-adjusted rate per 1,000 from 2016-2018 of county residents aged 65+ for whom a cardiovascular disease was the principle (or first-listed) diagnosis upon admission to the hospital [10].
6. *The United States Diabetes Surveillance System* under the guidance of the Division of Diabetes Translation, CDC asked adults if “a doctor has ever told you that you have diabetes” to calculate the percentage of adults who had diabetes per county. They also determined the percentage of adult that were obese, or whose body mass index (BMI) was over 30 from self-reported height and weight. The percentage of physically inactive was calculated from the number of adults who answered that they had not participated in physical activity in the last month [28].
7. The rate of active doctors, including MD’s and DO’s, per county from 2019-2020 was provided by the *Heath Resources and Services Administration* via the Area Heath Resource Files [17].

The graphical summaries of some of the socioeconomic factors considered in this paper are presented in Figure 8.

**Figure 8.:**
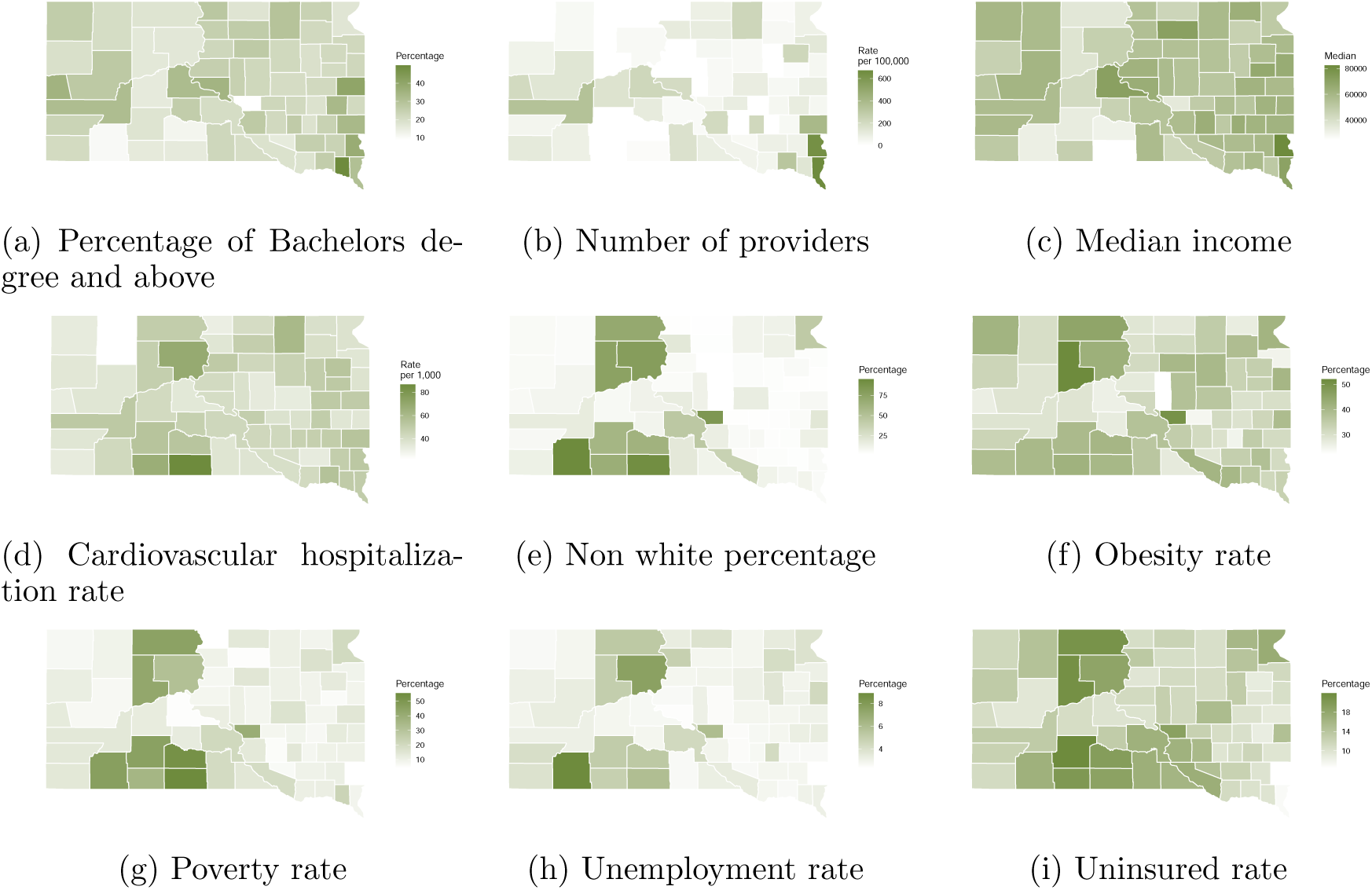
Graphical summary of the socioeconomic factors considered in this study.

### 6.2. Analysis for counties with 10 largest cities in SD

In SD, only a few of the counties had implemented any of the recommended mitigation strategies such as masking at the county level. In this section, some results are presented for the top ten counties in SD. These counties are chosen based on the population of the main city in that county. These top 10 counties with the largest cities in order of their size are: Minnehaha (Sioux Falls), Pennington (Rapid City), Brown (Aberdeen), Brookings (Brookings), Codington (Watertown), Davison (Mitchell), Yankton (Yankton), Hughes (Pierre), Beadle (Huron), and Lawrence (Spearfish) county. Several of counties are also home to South Dakota Board of Regent Schools. In order of size, South Dakota State University (Brookings), University of South Dakota (Clay), Black Hills State University (Lawrence), Northern State University (Brown), and South Dakota School of Mines and Technology (Pennington). This study took a closer look at the Brookings County where the South Dakota State University (SDSU) is located. This county is the first to put in place some mitigation strategies before any other county in the state.

Figure 9(a) shows the cumulative hospitalizations for the top ten counties. Minnehaha and Brown had the highest rates. Minnehaha had 932 hospitalizations per 100,000. In contrast, Brookings, Beadle, and Lawrence had the lowest rates. Brookings had 510 hospitalizations per 100,000 and Lawrence had 324 hospitalizations per 100,000. Figure 9(b) shows Davison and Codington had the highest death rates. Davison had 247 deaths per 100,000. Lawrence, Brookings, and Yankton had the lowest death rates. Brookings had 137 deaths per 100,000 and Yankton had 120 deaths per 100,000.

**Figure 9.:**
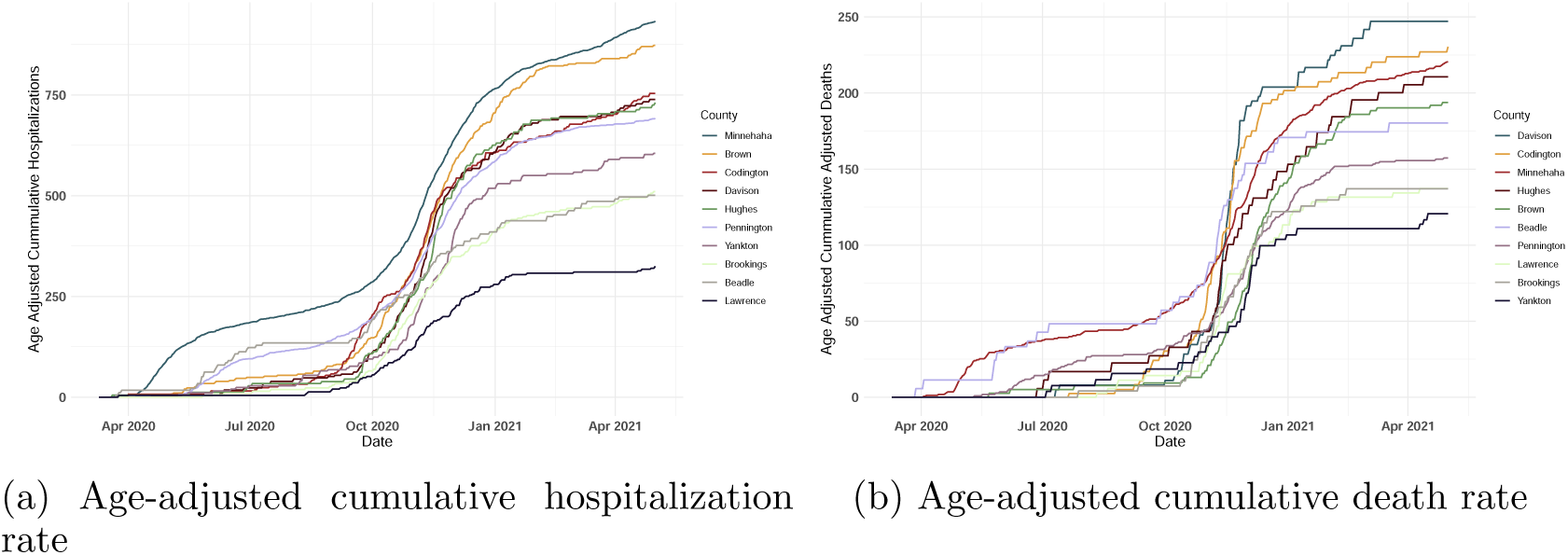
Trend of cumulative age-adjusted rates in the top 10 counties in South Dakota.

We compared the SIR values for the 10 counties (See Figure 10). For hospitalizations, Lawrence, Clay, Beadle, and Brookings had the lowest *SIR* values with Lawrence County experiencing about 60 percent fewer hospitalizations and Brookings experiencing about 36 percent fewer hospitalizations than expected according to the state rates. Minnehaha had 17 percent more and Brown had 10 percent more hospitalizations than expected. For deaths, Yankton, Brookings, Clay, and Lawrence Counties had the lowest *SIR* values with Yankton experiencing about 43 percent fewer deaths and the others about 35 percent fewer deaths. Only Davison County had significantly more deaths than expected, at about 17 percent more.

**Figure 10.:**
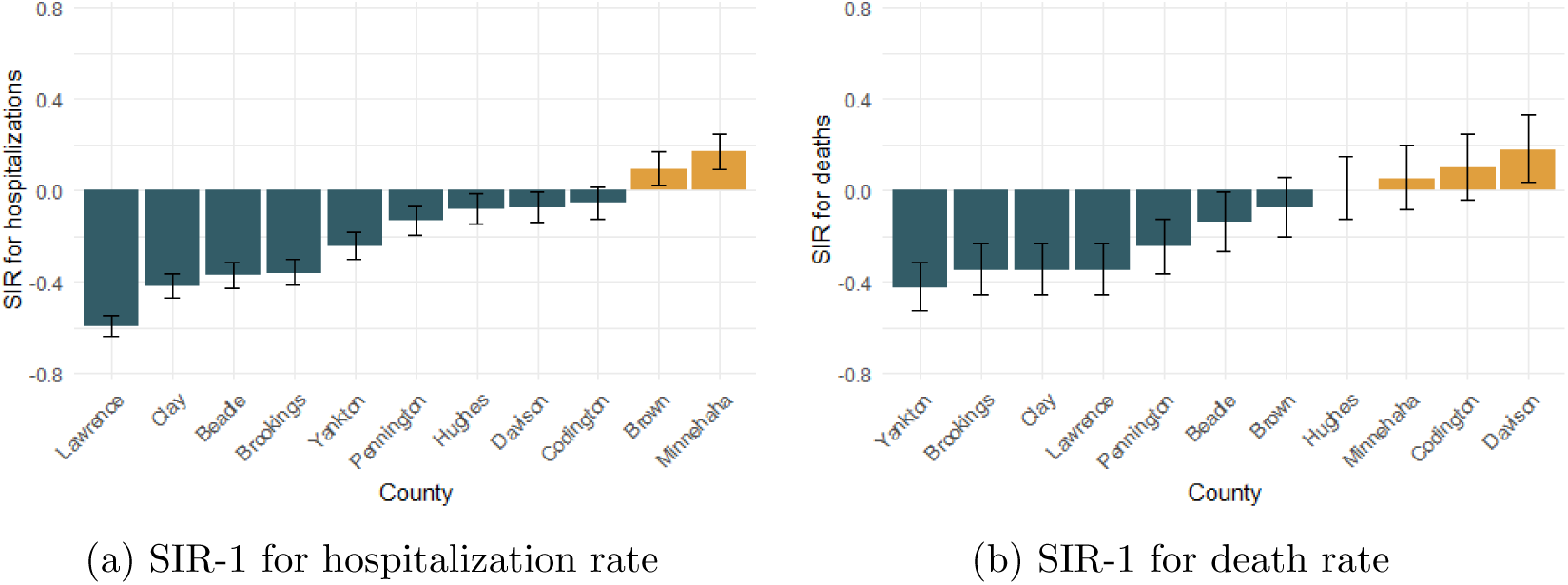
SIR-1 values and corresponding confidence bounds for the 10 counties in South Dakota.

In comparison to other counties in South Dakota, Brookings had a low uninsured percentage and nonwhite population. In contrast, the county experienced a higher rate of high educational attainment. In addition, Brookings county is generally a younger population, but this was controlled for using age-adjustments. In the scope of the COVID-19 pandemic, the city of Brookings’ local government, in Brookings County, was the first to take mitigating action against the disease, including limited capacity at public venues, enhanced cleaning and disinfecting guidelines, social distancing recommendations, and an enforceable mask mandate. In addition to balancing the health needs of the local population in Brookings county, the influx of young adults attending South Dakota State University in the city of Brookings, also posed additional considerations while navigating the pandemic. Through all of this, Brookings still fared well when compared to other comparable counties in South Dakota.

